# Structural synaptic signatures of Alzheimer’s Disease and Dementia with Lewy Bodies in the male brain

**DOI:** 10.1101/2021.09.30.21264355

**Authors:** Oleg O. Glebov, David Williamson, Dylan M. Owen, Tibor Hortobágyi, Claire Troakes, Dag Aarsland

**Affiliations:** Institute of Neuroregeneration and Neurorehabilitation, Qingdao University, Qingdao 266071, Shandong, China; Department of Old Age Psychiatry, The Institute of Psychiatry, Psychology & Neuroscience, King’s College London, De Crespigny Park, Denmark Hill, London SE5 8AF, UK; Randall Centre for Cell and Molecular Biophysics, Faculty of Life Sciences and Medicine, King’s College London, London SE1 1UL, UK; Institute of Immunology and Immunotherapy, School of Mathematics and Centre of Membrane Proteins and Receptors (COMPARE), University of Birmingham, Birmingham, UK; ELKH-DE Cerebrovascular and Neurodegenerative Research Group and Department of Neurology, Faculty of Medicine, University of Debrecen, Debrecen, Hungary; London Neurodegenerative Diseases Brain Bank, Department of Basic and Clinical Neuroscience, Institute of Psychiatry, Psychology and Neuroscience, King’s College London, London SE5 8AF, UK; Centre for Age-Related Medicine (SESAM), Stavanger University Hospital. Stavanger, Norway

## Abstract

The impact of Alzheimer’s Disease (AD) and Dementia with Lewy Bodies (DLB) on synaptic organisation remains poorly understood. Here, we found that in humans, DLB and AD were associated with increased synaptic levels of glutamate transporter vGlut1 and active zone protein Bassoon clustering respectively; these effects were only observed in male brain samples. These findings demonstrate disease- and sex-specific presynaptic structural remodelling in age-related neurodegenerative disorders.

## Main text

Alzheimer’s Disease (AD) and Dementia with Lewy Bodies (DLB) are the two major forms of neurodegenerative dementia, characterised by distinct pathological hallmarks involving aggregation of misfolded proteins. One proposed key pathophysiological mechanism in AD and DLB is synaptic dysfunction^1–6^; however, human synaptic pathology in AD and DLB besides synaptic loss is not characterised, and the underlying processes remain unknown.

Our previous studies identified distinct patterns of aberrant protein expression in AD and DLB^5,7^, raising the possibility that manifestation of AD and DLB may differ at the local synaptic level. To directly address this, we employed immunostaining, confocal and super-resolution microscopy to visualise synaptic structure in post mortem human brain isolates (**Fig. S1**) from Brodmann Area 9 (BA9), using a cohort of 32 control subjects and cases with severe AD and DLB.

We first assessed synaptic structure in 24 cases (**Table 1.1, Fig. S2**) by immunostaining for presynaptic active zone protein Basson (Bsn) and postsynaptic density protein Homer. The Homer/Bsn ratio showed no correlation with the post mortem interval (PMI), suggesting that synaptic structure was largely unaffected by preparation and storage (**Figure S2c**). There were no significant differences in Bsn levels, median synapse size as measured by Bsn, as well as RIM and Homer levels (**Figure 1a, S3a-c, S3e**), indicating that AD or DLB did not majorly alter the general morphology of the AZ and PSD.

**Table 1.**
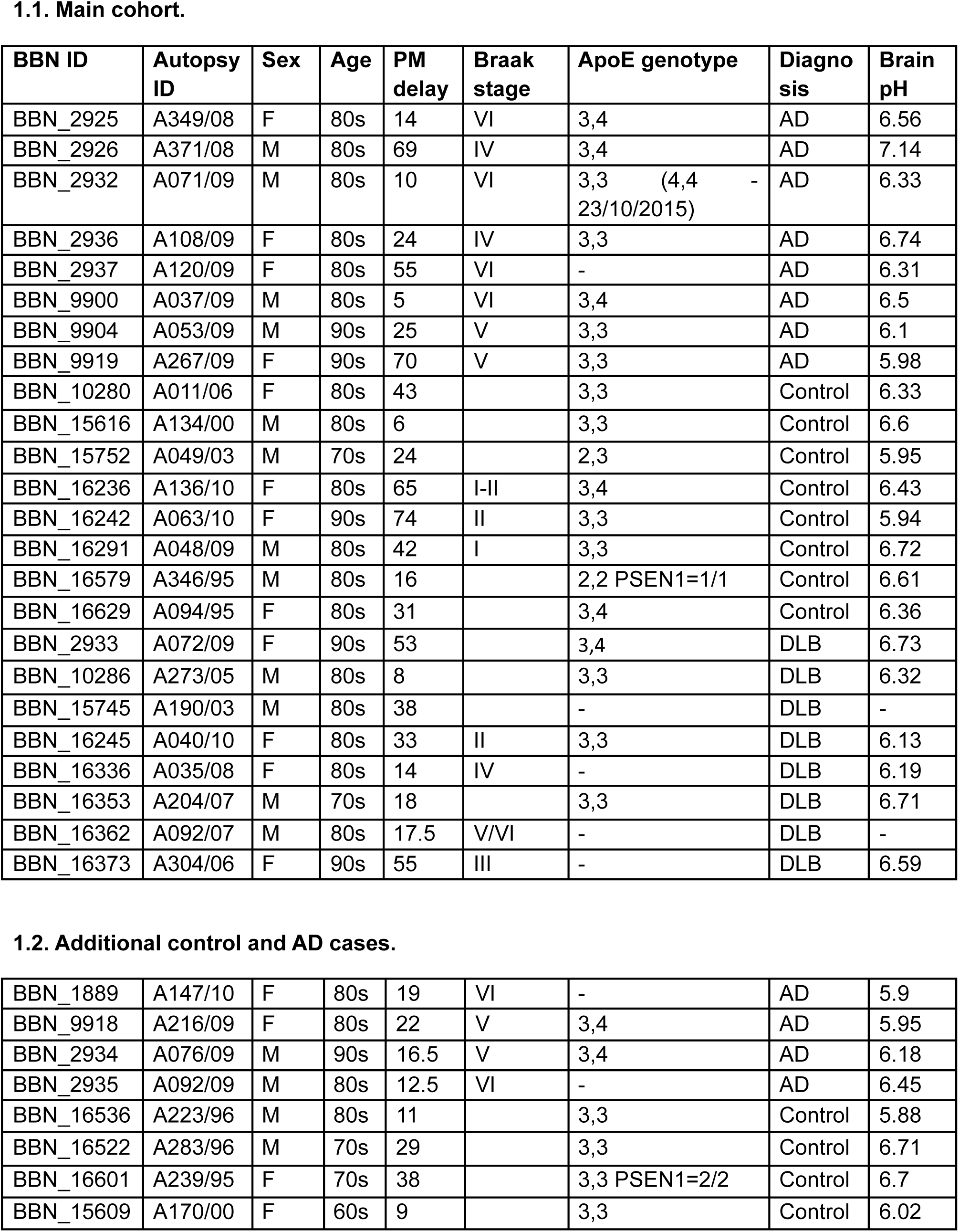
Summary of clinical cases.

**1.1. Main cohort.**
| BBN ID | Autopsy ID | Sex | Age | PM delay | Braak stage | ApoE genotype | Diagnosis | Brain pH |
| --- | --- | --- | --- | --- | --- | --- | --- | --- |
| BBN_2925 | A349/08 | F | 80s | 14 | VI | 3,4 | AD | 6.56 |
| BBN_2926 | A371/08 | M | 80s | 69 | IV | 3,4 | AD | 7.14 |
| BBN_2932 | A071/09 | M | 80s | 10 | VI | 3,3 (4,4 - 23/10/2015) | AD | 6.33 |
| BBN_2936 | A108/09 | F | 80s | 24 | IV | 3,3 | AD | 6.74 |
| BBN_2937 | A120/09 | F | 80s | 55 | VI | - | AD | 6.31 |
| BBN_9900 | A037/09 | M | 80s | 5 | VI | 3,4 | AD | 6.5 |
| BBN_9904 | A053/09 | M | 90s | 25 | V | 3,3 | AD | 6.1 |
| BBN_9919 | A267/09 | F | 90s | 70 | V | 3,3 | AD | 5.98 |
| BBN_10280 | A011/06 | F | 80s | 43 |  | 3,3 | Control | 6.33 |
| BBN_15616 | A134/00 | M | 80s | 6 |  | 3,3 | Control | 6.6 |
| BBN_15752 | A049/03 | M | 70s | 24 |  | 2,3 | Control | 5.95 |
| BBN_16236 | A136/10 | F | 80s | 65 | I-II | 3,4 | Control | 6.43 |
| BBN_16242 | A063/10 | F | 90s | 74 | II | 3,3 | Control | 5.94 |
| BBN_16291 | A048/09 | M | 80s | 42 | I | 3,3 | Control | 6.72 |
| BBN_16579 | A346/95 | M | 80s | 16 |  | 2,2 PSEN1=1/1 | Control | 6.61 |
| BBN_16629 | A094/95 | F | 80s | 31 |  | 3,4 | Control | 6.36 |
| BBN_2933 | A072/09 | F | 90s | 53 |  | 3,4 | DLB | 6.73 |
| BBN_10286 | A273/05 | M | 80s | 8 |  | 3,3 | DLB | 6.32 |
| BBN_15745 | A190/03 | M | 80s | 38 |  | - | DLB | - |
| BBN_16245 | A040/10 | F | 80s | 33 | II | 3,3 | DLB | 6.13 |
| BBN_16336 | A035/08 | F | 80s | 14 | IV | - | DLB | 6.19 |
| BBN_16353 | A204/07 | M | 70s | 18 |  | 3,3 | DLB | 6.71 |
| BBN_16362 | A092/07 | M | 80s | 17.5 | V/VI | - | DLB | - |
| BBN_16373 | A304/06 | F | 90s | 55 | III | - | DLB | 6.59 |
| **1.2. Additional control and AD cases.** |  |  |  |  |  |  |  |  |
| BBN_1889 | A147/10 | F | 80s | 19 | VI | - | AD | 5.9 |
| BBN_9918 | A216/09 | F | 80s | 22 | V | 3,4 | AD | 5.95 |
| BBN_2934 | A076/09 | M | 90s | 16.5 | V | 3,4 | AD | 6.18 |
| BBN_2935 | A092/09 | M | 80s | 12.5 | VI | - | AD | 6.45 |
| BBN_16536 | A223/96 | M | 80s | 11 |  | 3,3 | Control | 5.88 |
| BBN_16522 | A283/96 | M | 70s | 29 |  | 3,3 | Control | 6.71 |
| BBN_16601 | A239/95 | F | 70s | 38 |  | 3,3 PSEN1=2/2 | Control | 6.7 |
| BBN_15609 | A170/00 | F | 60s | 9 |  | 3,3 | Control | 6.02 |

**Figure 1.**
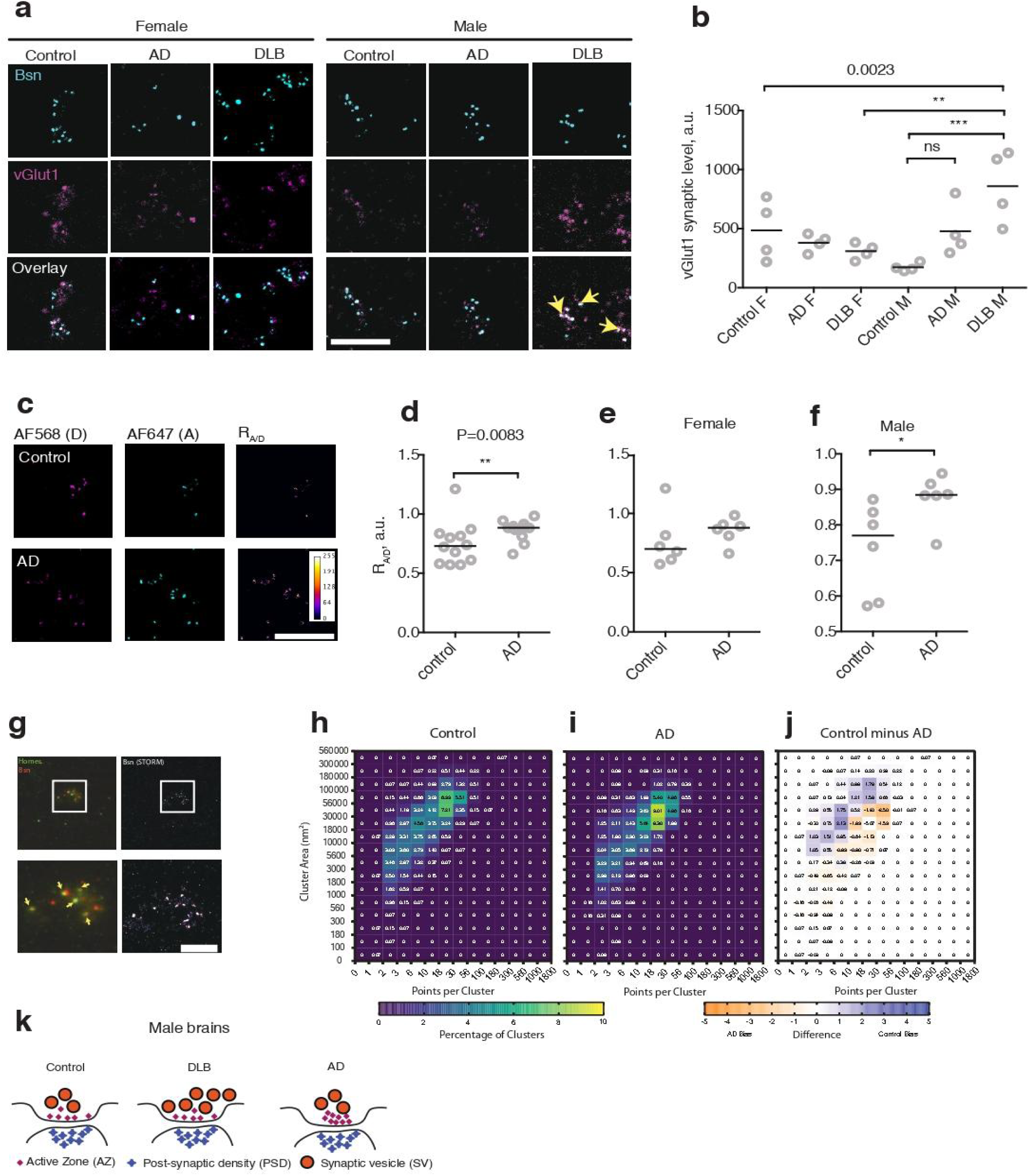
Disease-specific structural signatures of dementia in the male brain. **a**, Representative confocal microscopy images of human *post mortem* synaptoneurosomes fixed and immunostained for Bsn (cyan) and vGlut1 (magenta). Scale bar, 10 μm. **b**, vGlut1 synaptic levels in control, AD and DLB samples from female (F) and male (M) brains. ***P<0.0001, **P<0.01, 1-way ANOVA and Holm-Šidák’s post-test. **c**, Representative images of neurosynaptosome samples labelled for a ratiometric Bsn clustering assay. Scale bar, 10 μm. **d**, Ratiometric clustering measured in control vs AD samples. **P<0.0083, Mann-Whitney test. **e**, Ratiometric clustering in samples from female brains shows no significant difference between control and AD. P=0.4399, t-test. **f**, Ratiometric clustering in male brains shows significant difference between control and AD. *P<0.05, t-test. **g**, Representative image of synaptoneurosomal preparation immunostained for Homer (Alexa Fluor 488, green) and Bsn (Alexa Fluor 647, red). Arrows show multiple instances of Bsn puncta overlaying or adjacent to Homer puncta, indicative of synaptic structures. The right panels show STORM image of the Bsn channel; scale bar, 5 μm. **h**, Area and size of Bsn clusters in control samples, expressed as a normalized 2D histogram heatmap for the control condition. **i**, Same for AD samples. **j**, The difference in relative distribution of clusters into the histogram bins between the Control and AD conditions. N=3036 and 4153 clusters for Control and AD respectively, 4 brains per condition. **k**, A putative model of disease-specific presynaptic remodeling in male BA9 synapses with AD and DLB. DLB results in increased recruitment of vGlut1. AD results in an increased density of AZ matrix clustering, while PSD remains relatively unaffected by either DLB or AD.

We then performed double immunostaining for Bsn and vGlut1, a key component of synaptic vesicles (SV) that has been implicated in AD, as well as in Parkinson’s disease (PD), which shares many neuropathological features of DLB. Synaptic vGlut1 was elevated in both AD and DLB male samples compared to the controls, but the increase in AD samples was not statistically significant. In contrast, synaptic vGlut1 was significantly increased in male DLB samples compared to male controls or female DLB samples (**Figure 1a,b, S3d**). Therefore, we conclude that vGlut1 is specifically enriched in synapses from male subjects with DLB.

Recent evidence shows that neuronal activity regulates nanoscale clustering of the AZ^8^; we therefore reasoned that aberrant synaptic activity in AD may lead to nanoscale changes in AZ architecture, and investigated this in an extended cohort including 24 control and AD female and male cases (**Table 1.2, Figure S4**). We assessed Bsn clustering using confocal microscopy and a ratiometric assay developed by us previously^8^ (**Figure S5a**). Clustering in AD samples was significantly increased in male but not female brains (**Figure 1c–f**), indicative of an increased density of Bsn packing within the AZ. Clustering did not significantly correlate with age (**Figure S4d**), suggesting that the difference between control and AD samples was not due to the difference in median age (**Figure S4a**).

Finally, we used super-resolution imaging and machine learning clustering analysis developed by ourselves^9^ to directly quantify Bassoon clustering in four control and four AD male brains. A decrease in median cluster area and an increase in the median number of clusters was observed (**Figure 1g–j**). However, nested two-tailed t-tests showed no significant difference between median values in control and AD samples (**Figure S5b,c**), indicative of considerable variability in super-resolution data obtained from *post mortem* human brain samples. Taken together, these data show that AD in the male brain is associated with nanoscale reorganisation of presynaptic architecture.

Our findings show that two major forms of dementia are associated with distinct changes in presynaptic organisation, providing direct evidence for disease-specific synaptic defects in neurodegeneration (**Figure 1k**); furthermore, we present evidence for sex-specific effects in dementia at the level of synaptic structure, in line with association between biological sex and clinical manifestation of dementia^10^. Some data from female samples showed similar trends to those observed in male samples, yet failed to reach the threshold of significance (**Figure 1e,f**), possibly reflecting the higher incidence of “pure” dementia in men versus a mixed LBD/AD pathology reported in women^11^. It remains to be determined whether the observed changes represent a direct effect of pathology or a form of compensatory synaptic plasticity^3^.

In order to enhance the exploratory power of the approach described in this paper, further investigation will require larger case cohorts combined with testing for multiple synaptic markers, with particular relevance for quantitative analysis of nanoscale synaptic structure. Thus, our initial observations reported here pave the way for deeper investigation of synaptic architecture in neurodegeneration, with the long-term potential for development of targeted diagnostics and therapies for dementia^3^. Last but not least, in-depth structural analysis of human synaptic dysregulation will allow for knowledge-based validation of animal models^12^, providing a much-needed boost for experimental investigation and therapeutic development in neurodegeneration.

## Materials and Methods

See Supplementary Information.

## Data Availability

The datasets generated during and/or analysed during the current study are available from the corresponding author on reasonable request.

## Data availability statement

The datasets generated and analysed during the current study are available from the corresponding author on reasonable request.

## Acknowledgments

The authors are grateful to M. Malcangio (Wolfson CARD, King’s College London) for provision of facilities. Super-resolution microscopy imaging was carried out in the Nikon Imaging Centre (King’s College London). O.O.G. is supported by The Lewy Body Society (OOG2019/2020) and Natural Science Foundation China (32070772). T.H. is supported by the Hungarian Brain Research Program (2017-1.2.1-NKP-2017-00002). D.W. and D.M.O. are supported by BBSRC (BB/R007365/1). The London Neurodegenerative Diseases Brain Bank at King’s College London receives funding from the MRC and as part of the Brains for Dementia Research project (jointly funded by the Alzheimer’s Society and Alzheimer’s Research UK).

## Author contributions

Project was conceived by O.O.G., T.H. and D.A. Experimental methodology was developed by O.O.G., D.W. and D.M.O. Experimental procedures were carried out by O.O.G. and D.W. Experimental data analysis and interpretation was performed by O.O.G., D.W. and D.M.O. Clinical data was analysed by O.O.G., T.H., C.T. and D.A. Manuscript was drafted by O.O.G. and written with contributions from all authors.

## Supplementary information

### Materials and Methods

#### Post mortem human brain samples

Freshly frozen brain samples were provided by the London Neurodegenerative Diseases Brain Bank (KCL), part of the UK Brain Banks Network. The project was approved by the ethics committee of the The Institute of Psychiatry, Psychology & Neuroscience, King’s College London. Written informed consent from the donors and/or their relatives as appropriate was obtained by the London Neurodegenerative Diseases Brain Bank. Eight AD, eight DLB and eight control cases were included, with further four control and four AD cases included for extended analysis of synaptic structure in AD. Selection and diagnostic procedures have been described elsewhere^5,7^. All cases were diagnosed clinically with AD or DLB and this was confirmed by detailed neuropathological examination. All AD cases were classified as severe (Braak stage IV-VI), and all DLB cases exhibited diffuse neocortical pathology. Control cases showed no clinical evidence of cognitive decline and demonstrated only mild age-related pathology (Braak stage no more than II) on neuropathological examination.

Further details for all cases are presented in **Table 1**.

#### Biochemical preparation of the brain tissue

Small quantities (20-50 mg) of tissue from the cortex (Brodmann Area 9) were excised using a scalpel and homogenised in a Dounce homogeniser for 20 strokes on ice in 300 µl of homogenisation buffer, consisting of phosphate buffered saline (PBS), phosphatase inhibitor cocktail, protease inhibitor cocktail, and 5 µM EDTA). The resulting suspension was transferred into 1.5 ml Eppendorf tubes and the large cell debris sediment was pelleted at 1000g at the benchtop microcentrifuge for 10 min at 4°C. The supernatants from all brain samples were diluted with the homogenisation buffer to the same concentration (corresponding to 0.05 mg of the original brain material per 1 µl of buffer), aliquoted into 20 µl aliquots and stored at -70°C.

#### Immunocytochemistry staining

Brain extracts (20 µl) were added to 35 mm Petri dishes containing 1.5 ml of 2% paraformaldehyde (PFA) in PBS and 2-3 12 mm glass coverslips (thickness 1.5) coated with poly-L-lysine; to pellet the fixed material, Petri dishes were then centrifuged in the cell culture benchtop centrifuge for 20 min at 2800g. Coverslips were washed 4 times in excess of PBS and permeabilized in 0.2% Triton-X100 in PBS supplemented with 5% horse serum for 10 min. For immunostaining, coverslips were transferred into individual wells of a 24-well plate. Subsequent incubations were carried out in the permeabilization buffer. Coverslips were incubated with appropriate primary antibodies (see **Table 2**) for 60 min at room temperature (RT), washed 4 times in PBS and incubated with Alexa Fluor 488 and Alexa Fluor 647 conjugated secondary antibodies as appropriate at a concentration of 0.2 μg/mL each for 60 min at RT. Coverslips were then washed 4 times in PBS, mounted in Fluoromount-G mounting medium (Southern) on microscopy slides and imaged on a Zeiss LSM710 microscope equipped with a standard set of lasers through a Plan-Apochromat 63x/1.4 Oil objective. The imaging system was controlled by ZEN software. Regions of interest sized 1024 by 1024 pixels (65.8 nm/pixel) were imaged at speed 7 with the averaging setting 2. Pinhole size was kept to 1-2 Airy units. Excitation laser wavelengths were 543 and 633nm. Bandpass filters were set at 570-650 nm (Alexa Fluor 488) and 650–750nm (Alexa Fluor 647). Image acquisition was carried out at the 12-bit rate. Settings were optimized to ensure appropriate dynamic range, low background and sufficient signal/noise ratio.

**Table 2.**
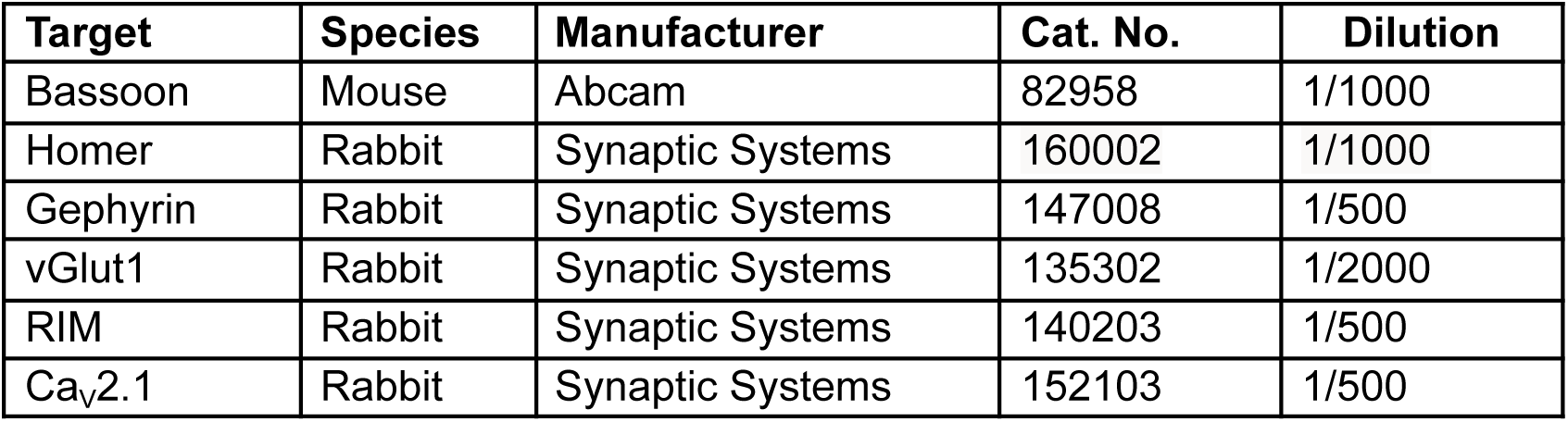
Primary antibodies used in the study.

| **Target** | **Species** | **Manufacturer** | **Cat. No.** | **Dilution** |
| --- | --- | --- | --- | --- |
| Bassoon | Mouse | Abcam | 82958 | 1/1000 |
| Homer | Rabbit | Synaptic Systems | 160002 | 1/1000 |
| Gephyrin | Rabbit | Synaptic Systems | 147008 | 1/500 |
| vGlut1 | Rabbit | Synaptic Systems | 135302 | 1/2000 |
| RIM | Rabbit | Synaptic Systems | 140203 | 1/500 |
| Ca<sub>v</sub>2.1 | Rabbit | Synaptic Systems | 152103 | 1/500 |

#### Ratiometric clustering assay

Coverslips were processed as described above for immunocytochemistry and incubated with a 1:1 mixture of donor- and acceptor-conjugated secondary antibodies at a concentration of 0.2 µg/ml each for 60min at RT. Alexa Fluor 568 and Alexa Fluor 647 were used as donor and acceptor respectively. Coverslips were then mounted in mounting medium (Southern), allowed to dry for 30min at RT and imaged on a Zeiss LSM710 microscope equipped with a standard set of lasers through a 63x oil objective. Excitation wavelengths were 543 and 633 nm. Bandpass filters were set at 560-615 (Alexa Fluor 568) and 650-750 nm (Alexa Fluor 647). Image acquisition was carried out as described above.

#### Confocal microscopy and image analysis

To identify individual synapses, images were thresholded in ImageJ using the “Moments” setting, resulting in binary images, and particles were then counted automatically using the “Analyze Particles” command across the whole image. Visible areas of non-specific autofluorescence associated with tissue debris were excluded from analysis. Thresholded data from the Bassoon (Bsn) channel was used to determine synaptic locations as Regions Of Interest (ROI). Signal intensities were quantified for each synaptic puncta using the ROI Manager function. To avoid rare overlap of multiple synapses, only ROIs with areas ranging from 0.1 to 2 μm^2^ were included in further analysis. All values of circularity were included in analysis. Mean intensities were then calculated for each synapse, and the median value describing the distribution of synaptic intensities in the whole sample was reported for further statistical analysis. Background subtraction was performed as required. To quantify synapse-specific ratios R_A/D_, puncta of fluorescence were individually processed in the same manner.

#### Validation of the synaptosomal preparation

Immunohistochemical analysis of synapses in intact brain structure is associated with considerable artifacts due to incomplete reagent penetration and signal density, complicating both conventional and super-resolution microscopy^13^. Therefore, we opted for biochemical purification of the synaptoneurosomal fraction (**Figure S1a**), which isolates structurally intact synapses and has been well established in structural and functional studies of brain synapses^14–16^. To label visualize synapses, we used immunostaining for Bsn (see main text).

Bsn-positive puncta colocalized with the key classes of synaptic markers; these included inhibitory postsynaptic density (PSD) protein Gephyrin, excitatory postsynaptic density protein Homer, presynaptic vesicle (SV) glutamate transporter vGlut1 and voltage-gated calcium channel (VGCC) Cav2.1 (**Figure S1b**). Furthermore, the median size of the synapse as evidenced by Bsn staining was not altered in the synaptoneurosomal preparation, suggesting that the protocol preserved synaptic structure, did not affect synaptic size and likely retained the structural diversity of the synaptic population (**Figure S1c**). In agreement with this, synapse-specific levels of Bsn were similar in the intact brain material and the synaptosomal preparation (**Figure S1d**). However, labelling for vGlut1 was drastically lower in the intact brain sample compared to the purified synaptosomal fraction, suggesting impaired penetration of the anti-vGlut1 antibody into the intact brain tissue (**Figure S1e**). Furthermore, coefficients of variation for vGlut1 and Bsn intensities in intact samples were considerable (72.36% and 55.85% respectively). Taken together, these data show that biochemically purified synaptoneurosomes enriched in the preparation are more amenable for immunostaining compared to intact brain samples, in agreement with previously published data^15,17^. Therefore, the above protocol was used for the rest of the study.

#### Super-resolution microscopy

Samples were fixed and stained with Alexa Fluor 647 as for confocal imaging, except brain extracts were deposited in glass-bottomed chamber slides (#1.5 glass, ibidi μSlides) coated with PLL. For imaging, the final PBS wash was replaced with a volume of freshly prepared STORM imaging buffer (50 mM Tris-HCI (pH 8.5), 10 mM NaCl, 0.56M glucose, 5 U/ml pyranose oxidase (Sigma P4234), 40 μg/ml bovine catalase (Sigma C40), 35 mM cysteamine (Sigma 30070), and 2 mM cyclooctatetraene (Sigma 138924)). The dSTORM image sequences were acquired on a Nikon N-STORM 5 system in a TIRF configuration using a CFI SR HP Apochromat TIRF 100×AC 1.49 NA oil objective, arranged for a pixel size of 160 nm. Samples were illuminated with 647 nm laser light at approximately 2.05 kW/cm^2^. Images were recorded using a Hamamatsu ORCA-Flash4.0 scientific CMOS camera using a centered 256 × 256 pixel region at 20 ms per frame for 15-20,000 frames.

#### Super-resolution microscopy data analysis

Data were processed using ThunderSTORM^18^ version ‘dev-2015-10-03-b1’ and the following parameters: pre-detection wavelet filter (B-spline, scale 2, order 3), initial detection by non-maximum suppression (radius 1, threshold at one standard deviation of the F1 wavelet), and sub-pixel localization by integrated Gaussian point-spread function (PSF) and maximum likelihood estimator with a fitting radius of 3 pixels. Detected points were corrected for sample drift using cross-correlation of images from 5 bins at a magnification of 5. The occurrence of repeated localizations, such as can occur from long dye on times or fast re-blinking, was reduced by merging points (in the drift-corrected dataset) which reappeared within 50 nm and 25 frames of the initial detection. For the purposes of interpretation, it is assumed that the frequency of multiple detection of dye molecules is independent of the sample staining and therefore the relative changes in the clustering of points between sample conditions are independent of dye re-blinking. The merged dataset was then filtered with points retained according to the following criteria: an intensity range of 450 - 10000 photons, a sigma range of 50 - 250, and a localization uncertainty of less than 25 nm. The final dataset was then exported to a comma-delimited text file.

Data were analysed with CAML, a machine learning tool for SMLM cluster analysis recently developed by us^9^. Two SMLM images were manually annotated to label points within synapse-like structures. These data were then used to train a model to identify points which are both clustered and in synapse-like structures. This model, designated ‘2XOGG3’, demonstrated 98% accuracy in identifying points within synapses in the test dataset. The model was then used to annotate the remaining SMLM images and extract quantitative information on the clusters within synapses.

#### Statistical analysis

All of the synapses automatically detected within these fields of view were included in the analysis. Statistical analysis was carried out using the Prism 6.0c software package (GraphPad Software). Data distributions were assessed for normality using d’Agostino and Pearson omnibus normality test. All tests were unpaired and two-tailed. For normally distributed datasets, Student’s t-test, 1-way ANOVA and Holm-Šidák’s post-test were used to assess statistical significance as appropriate; for not normally distributed datasets, Mann-Whitney rank test, Kruskal-Wallis test and Dunn’s post-test were used for assessing statistical significance as appropriate. Datasets were presented as scatter dot plots with line at median or as cumulative probability plots, with error bars showing interquartile ranges where appropriate.

## Supplementary Figures

**Figure S1.**
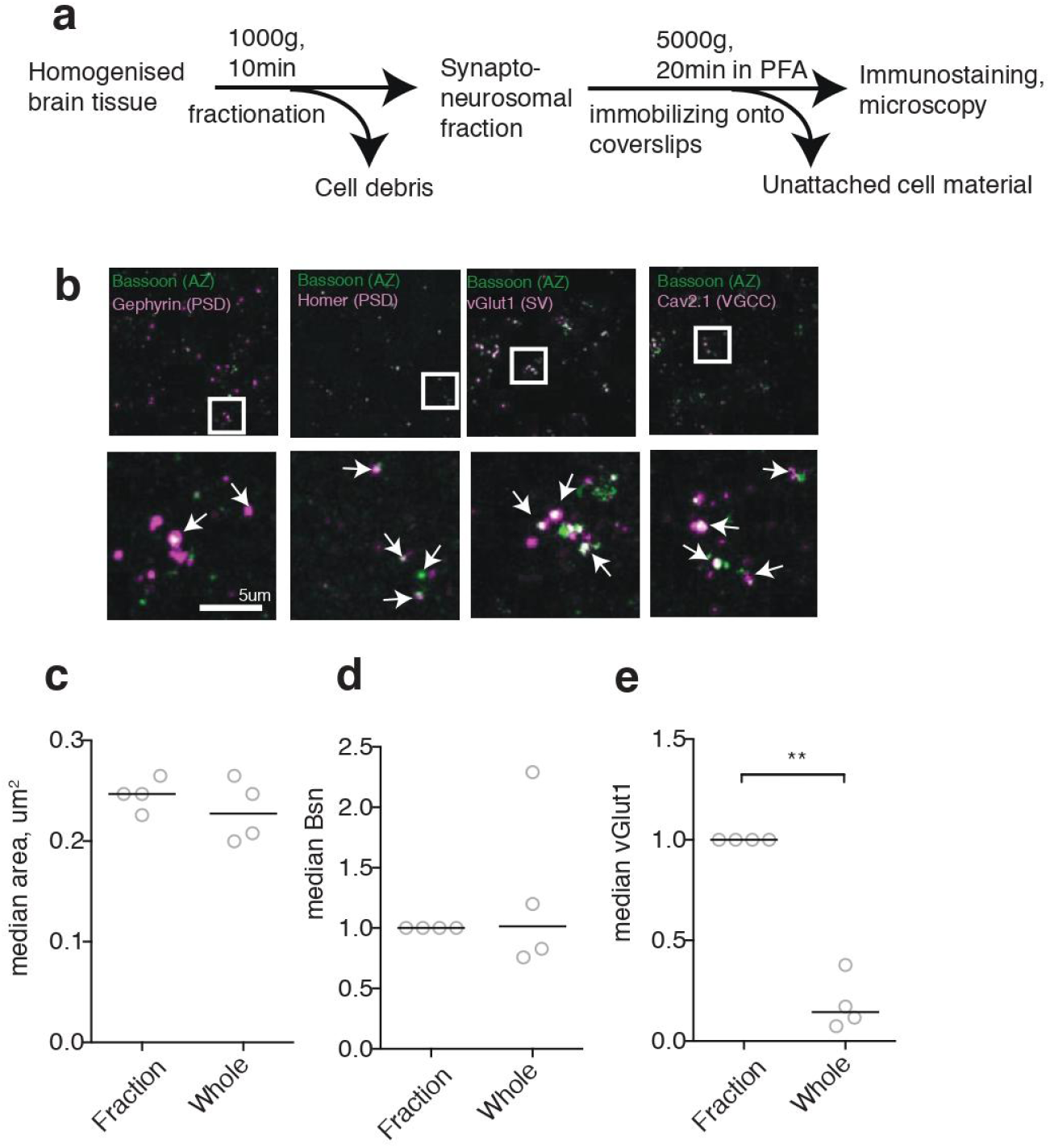
Characterization of the synaptic preparation. **a**, Schematics of the preparation of neurosynaptosomes. **b**, Colocalization between synaptic markers in neurosynaptosomes. **c**, Median synaptic area in neurosynaptosomal fraction (Fraction) and non-homogenised samples (Whole) from 4 brains. P=0.3881, t-test. **d**, Synaptic Bsn labelling in neurosynaptosomal fraction and non-homogenised samples from 4 brains. Intensities were normalised to neurosynaptosomal fraction. P=0.5036, one sample t-test. **e**, Synaptic vGlut1 labelling in neurosynaptosomal and non-homogenised samples from 4 brains. Intensities were normalised to neurosynaptosomal fraction. **P=0.0012, one sample t-test.

**Figure S2.**
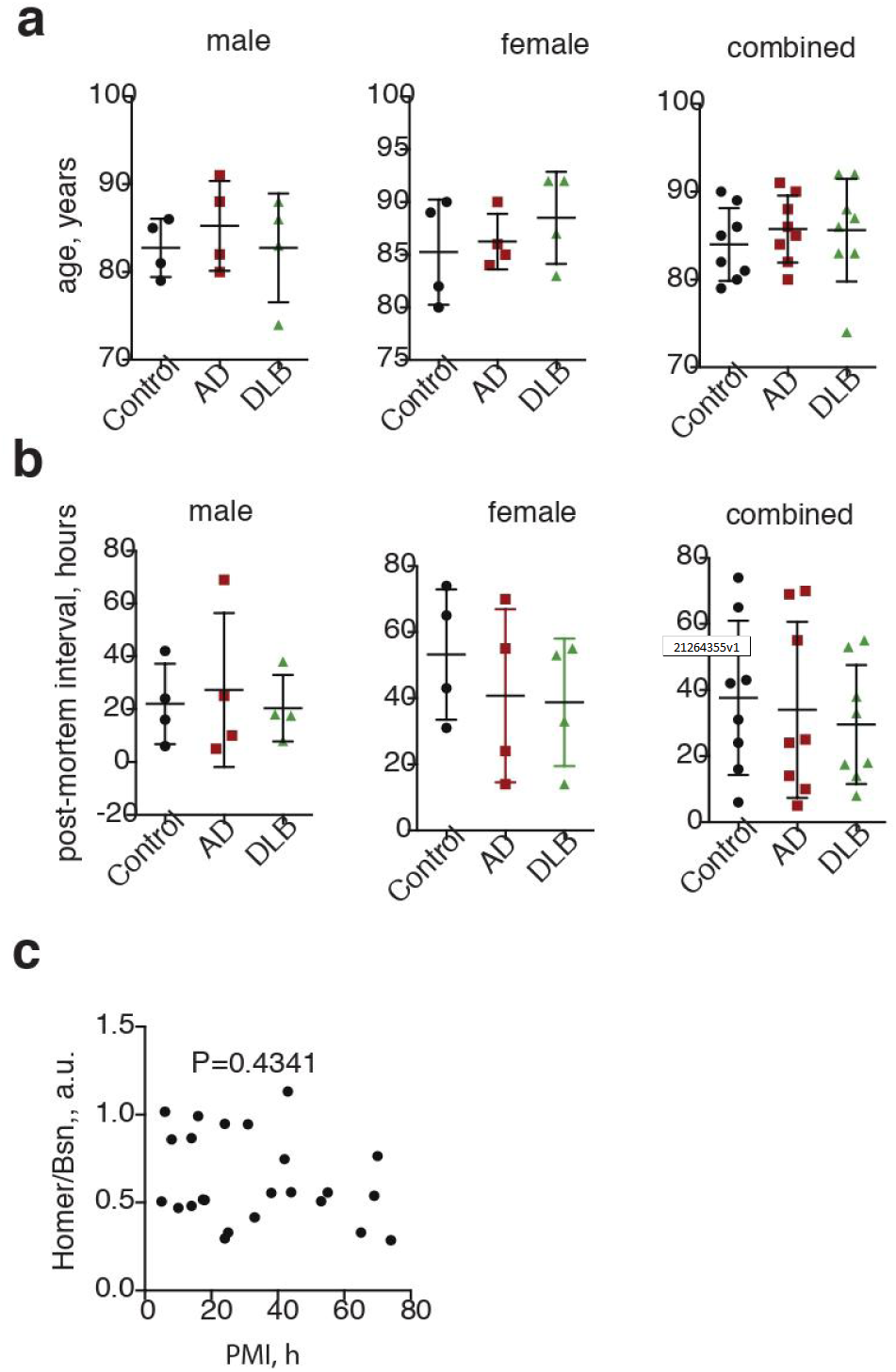
Supporting data for Table 1.1. **a**, Age is not significantly different between groups. P=0.7263 (male), P=0.5430 (female), P=0.7097 (both), 1-way ANOVA. **b**, PMI is not significantly different between groups P=0.8849 (male), 0.6154 (female), 0.7826 (both), 1-way ANOVA. **c**, Postsynaptic/presynaptic ratio does not correlate with PMI. P=0.4341, r=-0.1675, Spearman’s correlation coefficient.

**Figure S3.**
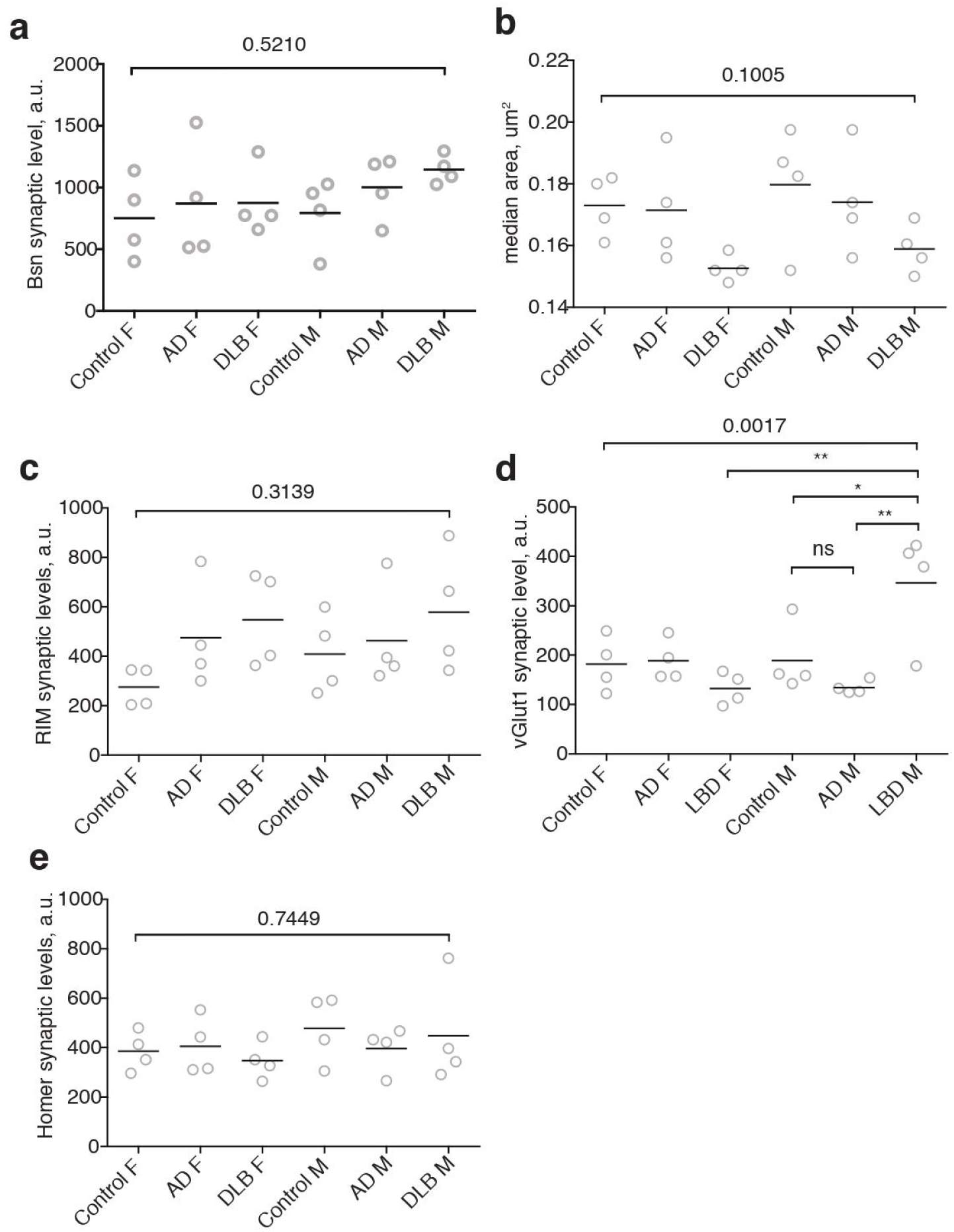
Supporting data for synaptic marker proteins levels 1. **a**, Bsn synaptic levels in control, AD and DLB samples from male and female brains P=0.5210, 1-way ANOVA. **b**, median synaptic area in samples from control, AD and DLB male and female brains P=0.1005, 1-way ANOVA. **c**, RIM synaptic levels in sample from control, AD and DLB female and male brains P=0.3139, 1-way ANOVA. **d**, vGlut1 synaptic levels in samples from control, AD and DLB female and male brains, second sample preparation. **P<0.01, *P<0.05, 1-way ANOVA and Holm-Šidák’s post-test. **e**, Homer synaptic levels in sample from control, AD and DLB female and male brains. P=0.7449, 1-way ANOVA.

**Figure S4.**
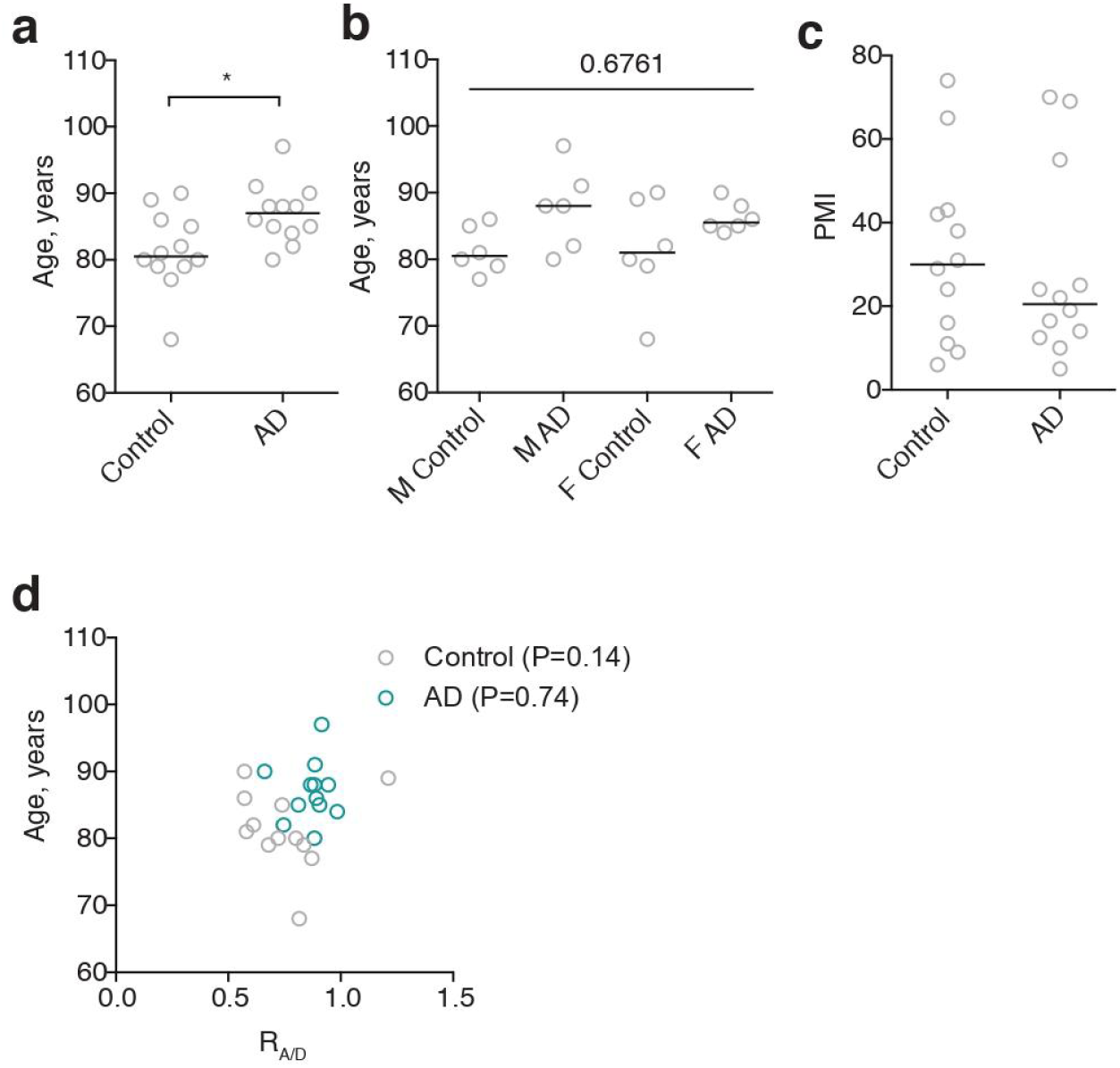
Supporting data for Table 1.2. **a**, Age of AD cases was significantly higher than that of control cases. *P<0.05, Student’s t test. **b**, 1-way ANOVA divided sex and condition shows no significant differences. P=0.1176, 1-way ANOVA. **c**, PMI is not significantly different. P=0.6761, t-test. **d**, R_A/D_ does not correlate with age of cases. P=0.1374 (Control), P=0.7429 (AD), r=-0.4526 (Control), r=0.1058 (AD), Spearman’s correlation coefficient.

**Figure S5.**
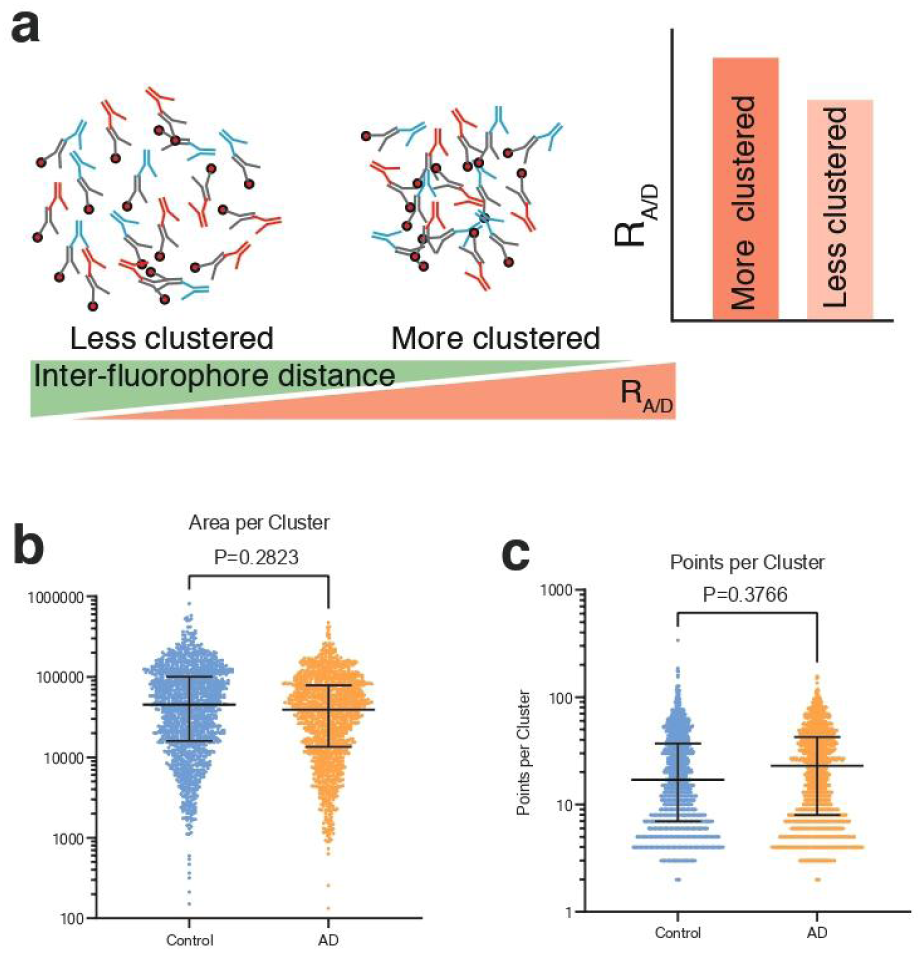
Supporting data for clustering experiments. **a**, Schematics of the ratiometric clustering assay – adapted from Ref. 8. **b**, Area of Bsn clusters in control and AD samples; pairwise comparison is a two-tailed nested t test. **c**, Localization counts for Bsn in control and AD samples; pairwise comparison is a two-tailed nested t test.

## References

1. Terry, R. D. et al. Physical basis of cognitive alterations in alzheimer’s disease: Synapse loss is the major correlate of cognitive impairment. Ann. Neurol. 30, 572–580 (1991).

2. Revuelta, G. J., Rosso, A. & Lippa, C. F. Neuritic Pathology as a Correlate of Synaptic Loss in Dementia With Lewy Bodies. Am. J. Alzheimers Dis. Dementiasr 23, 97–102 (2008).

3. Jackson, J. et al. Targeting the Synapse in Alzheimer’s Disease. Front. Neurosci. 13, 735 (2019).

4. Sheng, M., Sabatini, B. L. & Südhof, T. C. Synapses and Alzheimer’s disease. Cold Spring Harb. Perspect. Biol. 4, (2012).

5. Bereczki, E. et al. Synaptic proteins predict cognitive decline in Alzheimer’s disease and Lewy body dementia. Alzheimers Dement. 12, 1149–1158 (2016).

6. Schulz-Schaeffer, W. J. The synaptic pathology of alpha-synuclein aggregation in dementia with Lewy bodies, Parkinson’s disease and Parkinson’s disease dementia. Acta Neuropathol. (Berl.) 120, 131–43 (2010).

7. Bereczki, E. et al. Synaptic markers of cognitive decline in neurodegenerative diseases: a proteomic approach. Brain J. Neurol. 141, 582–595 (2018).

8. Glebov, O. O. et al. Nanoscale Structural Plasticity of the Active Zone Matrix Modulates Presynaptic Function. Cell Rep. 18, 2715–2728 (2017).

9. Williamson, D. J. et al. Machine learning for cluster analysis of localization microscopy data. Nat. Commun. 11, 1–10 (2020).

10. Podcasy, J. L. & Epperson, C. N. Considering sex and gender in Alzheimer disease and other dementias. Dialogues Clin. Neurosci. 18, 437–446 (2016).

11. Barnes, L. L., Lamar, M. & Schneider, J. A. Sex differences in mixed neuropathologies in community-dwelling older adults. Brain Res. 1719, 11–16 (2019).

12. King, A. The search for better animal models of Alzheimer’s disease. Nature 559, S13–S15 (2018).

13. Melvin, N. R. & Sutherland, R. J. Quantitative caveats of standard immunohistochemical procedures: Implications for optical disector-based designs. J. Histochem. Cytochem. 58, 577–584 (2010).

14. Hardy, J. A. et al. Use of post-mortem human synaptosomes for studies of metabolism and transmitter amino acid release. Neurosci. Lett. 33, 317–322 (1982).

15. Jhou, J.-F. & Tai, H.-C. The Study of Postmortem Human Synaptosomes for Understanding Alzheimer’s Disease and Other Neurological Disorders: A Review. Neurol. Ther. 6, 57–68 (2017).

16. Biesemann, C. et al. Proteomic screening of glutamatergic mouse brain synaptosomes isolated by fluorescence activated sorting. EMBO J. 33, 157–70 (2014).

17. Postupna, N. O. et al. Flow cytometry analysis of synaptosomes from post-mortem human brain reveals changes specific to Lewy body and Alzheimer’s disease. Lab. Investig. J. Tech. Methods Pathol. 94, 1161–72 (2014).

18. Ovesný, M., Krížek, P., Borkovec, J., Švindrych, Z. & Hagen, G. M. ThunderSTORM: A comprehensive ImageJ plug-in for PALM and STORM data analysis and super-resolution imaging. Bioinformatics 30, 2389–2390 (2014).

